# Association between Remnant Cholesterol and All-Cause and Cardiovascular Mortality Risk in Chronic Kidney Disease Patients Over 60: A Cohort Study Based on NHANES 2001-2018

**DOI:** 10.1101/2025.02.12.25322142

**Authors:** Jiechun YAO, Chun Lin, Qingbo Xu, Guode Li, Yan Lin

## Abstract

**Objective:** Chronic kidney disease (CKD) patients face significant mortality risks, and remnant cholesterol (RC) may be a critical prognostic indicator. This study aimed to explore the association between RC and all-cause and cardiovascular mortality risks in CKD patients.

**Methods:** This prospective cohort study utilized data from the National Health and Nutrition Examination Survey (NHANES) 2001-2018, including 1,606 CKD patients aged over 60. Cox proportional hazards models were employed to assess the association between RC and all-cause and cardiovascular mortality, with multi-model adjustments and subgroup analyses.

**Results:** During 93.9 months of follow-up, 932 all-cause mortality events (58.0%) and 283 cardiovascular mortality events (17.6%) were observed. Each logarithmic unit increase in RC was associated with a significant 41% reduction in all-cause mortality risk (adjusted HR: 0.59, 95% CI: 0.40-0.87, P=0.0079). Subgroup analyses revealed significantly reduced all-cause mortality risks in women (HR=0.71), hypertensive patients (HR=0.75), and type 2 diabetes patients (HR=0.66).

**Conclusion:** In CKD patients, RC is significantly associated with reduced all-cause mortality risk, providing a new perspective for individualized mortality risk assessment.

## Introduction

Chronic Kidney Disease (CKD) is a global public health concern, especially among individuals over 60(1). Recent studies show a CKD prevalence of 13.4% worldwide, with rates in China between 11.3% and 13.7% (2, 3). CKD patients face alarming outcomes, with all-cause mortality rates up to 40% and cardiovascular events as a primary cause of death(4). Cardiovascular disease incidence is significantly higher in CKD patients, particularly among the elderly(5).

Remnant Cholesterol (RC) represents cholesterol remnants remaining after triglyceride-rich lipoprotein metabolism(6). Increasingly recognized for cardiovascular risk, recent literature links elevated RC levels to adverse outcomes like atherosclerosis, heart attacks, and strokes(7, 8). In CKD patients, high RC levels are a critical marker for increased atherosclerotic cardiovascular disease (ASCVD) risk(9). However, the RC-disease relationship remains complex, with studies showing inconsistent associations(10).

The relationship between RC and mortality in CKD patients remains controversial. Supporting studies correlate higher RC levels with increased cardiovascular and all-cause mortality, particularly in kidney transplant recipients and diabetic CKD patients(11). Conversely, some research suggests varying mortality impacts across different disease states, with heart failure patients showing different RC-mortality relationships(12). While most studies suggest a positive correlation between high RC levels and increased mortality in CKD patients, conflicting results underscore the need for further investigation.

This study employed a prospective cohort research method, analyzing data from the NHANES database between 2001-2018 to evaluate the association between red cell distribution width RC and all-cause and cardiovascular mortality rates among CKD patients aged 60 and above.

## Methods

### Data and study participants

The research data were sourced from the National Health and Nutrition Examination Survey (NHANES,https://www.cdc.gov/nchs/nhanes/), an authoritative public health database managed by the National Center for Health Statistics (NCHS). NHANES ensures scientific rigor and representativeness through complex stratified and multi-stage sampling techniques(13, 14). The database comprehensively captures demographic characteristics, physical assessments, biochemical markers, and detailed dietary and health history, providing an invaluable resource for investigating the health status and lifestyle of the U.S. population.The study strictly adhered to the ethical guidelines of the 1975 Helsinki Declaration and obtained approval from the NCHS Research Ethics Review Committee. All participants provided voluntary informed consent, thereby safeguarding research ethics and participant rights.

This study systematically collected data covering 91,351 participants between 2001 and 2018. Initially, individuals under 60 years old (n=74,098) were excluded, followed by the removal of individuals lacking relevant RC data (n=12,841) and those with missing CKD data or classified as non-CKD (n=2,804). Subsequently, we further excluded individuals without all-cause mortality information (n=2). After this rigorous screening process, 1,606 participants ultimately met the inclusion criteria for this study, as illustrated in Fig. 1.

**Fig 1.**
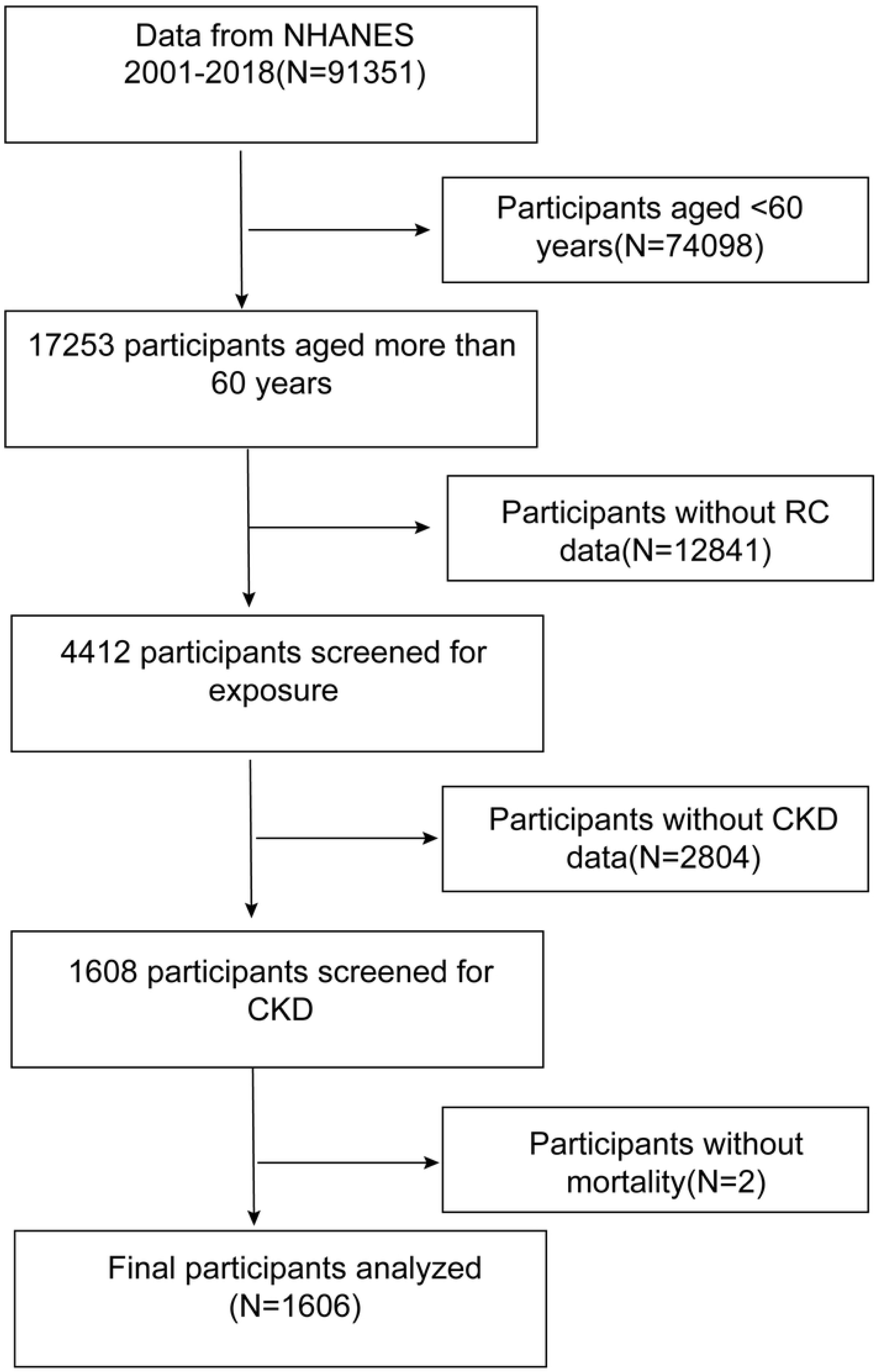
A flow chart of the sample selection from NHANES 2001–2018.RC, Remnant Cholesterol; CKD, chronic kidney disease.

### Exposed Variables and Outcomes

RC is a critical blood lipid assessment indicator calculated from a standard lipid profile in the fasting state. The calculation method is:

RC = Total Cholesterol (TC) - Low-Density Lipoprotein Cholesterol (LDL-C) - High-Density Lipoprotein Cholesterol (HDL-C)(15–17).

The primary study endpoints were all-cause and cardiovascular-specific mortality. We employed advanced data linkage techniques to match NHANES participant records with the National Death Index, tracking participant vital status from the survey date until December 31, 2006. Mortality classification followed the International Classification of Diseases, 10th Revision (ICD-10), with specific focus on cardiovascular mortality categories (I00-I09, I11, I13, I20-I51, I60-I69). Participants without matched death records were considered alive at the end of the follow-up period(18, 19).

### Covariates

We comprehensively assessed participants’ health status using standardized diagnostic criteria and validated assessment tools. Hypertension was defined by three criteria: ongoing antihypertensive medication, physician-confirmed diagnosis, or persistent blood pressure measurements ≥140/90 mmHg across three consecutive readings. Diabetes mellitus was diagnosed through multiple parameters, including antidiabetic medication use, physician diagnosis, or meeting specific laboratory thresholds: glycated hemoglobin A1c ≥6.5%, fasting blood glucose ≥126 mg/dL, or 2-hour postprandial glucose ≥200 mg/dL(20). The diagnosis of coronary artery disease (CAD) was determined through self-reported physician diagnoses obtained during personal interviews using a standardized medical condition questionnaire. Subjects were asked, “Has a doctor or other health professional ever told you that you have congestive heart failure/coronary heart disease/angina pectoris/heart attack/stroke?” Answering yes to any of these questions was coded as positive for coronary artery disease (CAD)(21).

Demographic and socioeconomic variables were meticulously documented to control for potential confounding factors. Racial categories included Mexican Americans, other Hispanic, non-Hispanic Black, non-Hispanic White, and other races (including multiracial). Education levels were stratified into five comprehensive categories ( less than 9th grade,9-11th grade,high school graduate/GED equivalent,some college or Associate’s degree and college graduate or above). Lifestyle characteristics were systematically assessed across three domains: smoking status (never smoked, current smokers, former smokers), alcohol consumption (drinkers versus non-drinkers), and anthropometric measurements, with body mass index (BMI) serving as a key indicator of physical health.

We employed rigorous biochemical analysis techniques to quantify key biomarkers with high precision. Serum and plasma creatinine concentrations were determined using the Jaffe rate method (kinetic alkaline picrate), a standard spectrophotometric technique known for its analytical accuracy. Urinary albumin was quantified through solid-phase fluorescence immunoassay (FIA), while urinary creatinine levels were measured using standardized enzymatic methods. Urine samples were collected randomly to minimize sampling bias and ensure representative measurement. The albumin-to-creatinine ratio (ACR) and estimated glomerular filtration rate (eGFR) were calculated using established mathematical formulas (22, 23), providing comprehensive insights into renal function and metabolic parameters.

ACR (mg/g) = urinary albumin(mg/dL)/urine creatinine(g/dL)

eGFR (ml/min/1.73m²) = 141.9 × (Scr/κ)^α^ × max(Scr/κ, 1)^-^¹·²⁰⁹ × 0.993ᴬᵍᵉ × 1.018(if female) × 1.159(if black)

It is important to note that Scr denotes serum creatinine concentration (mg/dL), and κ is 0.9 for males and 0.7 for females,and α is −0.411 for males and −0.329 for females.

The diagnosis of CKD is based on the following criteria(24–26): eGFR < 60 ml/min/1.73 m2 or ACR > 30 mg/g. Additionally, the study included various blood markers such as triglycerides (TG) (mg/dL), total cholesterol (TC) (mg/dL), high-density lipoprotein cholesterol (HDL-C) (mg/dL), low-density lipoprotein cholesterol (LDL-C) (mg/dL), serum creatinine (μmol/L), fasting blood glucose (mg/dL), and glycated hemoglobin (HbA1C (%)).

### Statistical analysis

The statistical analyses of this study were performed EmpowerStats (http://www.empowerstats.com, X&Y Solutions, Inc., Boston, MA).In this study, to enhance the representativeness of results and reduce potential bias, researchers used sampling weights as an important supplement to the analytical method. Continuous variables were presented as mean values with 95% confidence intervals, while categorical variables were described by occurrence frequency and corresponding percentages, and statistical significance was calculated using weighted χ2 tests. To investigate the relationship between RC and all-cause and cardiovascular mortality, we employed a weighted Cox regression model for survival analysis of the study population, using hazard ratios (HR) with 95% confidence intervals (CI) to assess the impact of RC on survival rates. We constructed three models, with the first model having no covariate adjustments; Model 2 incorporated adjustments for factors such as age, sex, and race; Model 3 added covariates from Table 1 on the basis of Model 2. Additionally, the dose-response effect between RC and all-cause and cardiovascular mortality was evaluated using a generalized additive model and fitted curves (using penalized spline method). Finally, to ensure the robustness of results, we conducted sensitivity analyses stratified by age (< 75 years or ≥ 75 years), sex, smoking status, hypertension, coronary heart disease, and diabetes. In all hypothesis tests, p-values < 0.05 were considered statistically significant.

**Table 1.**
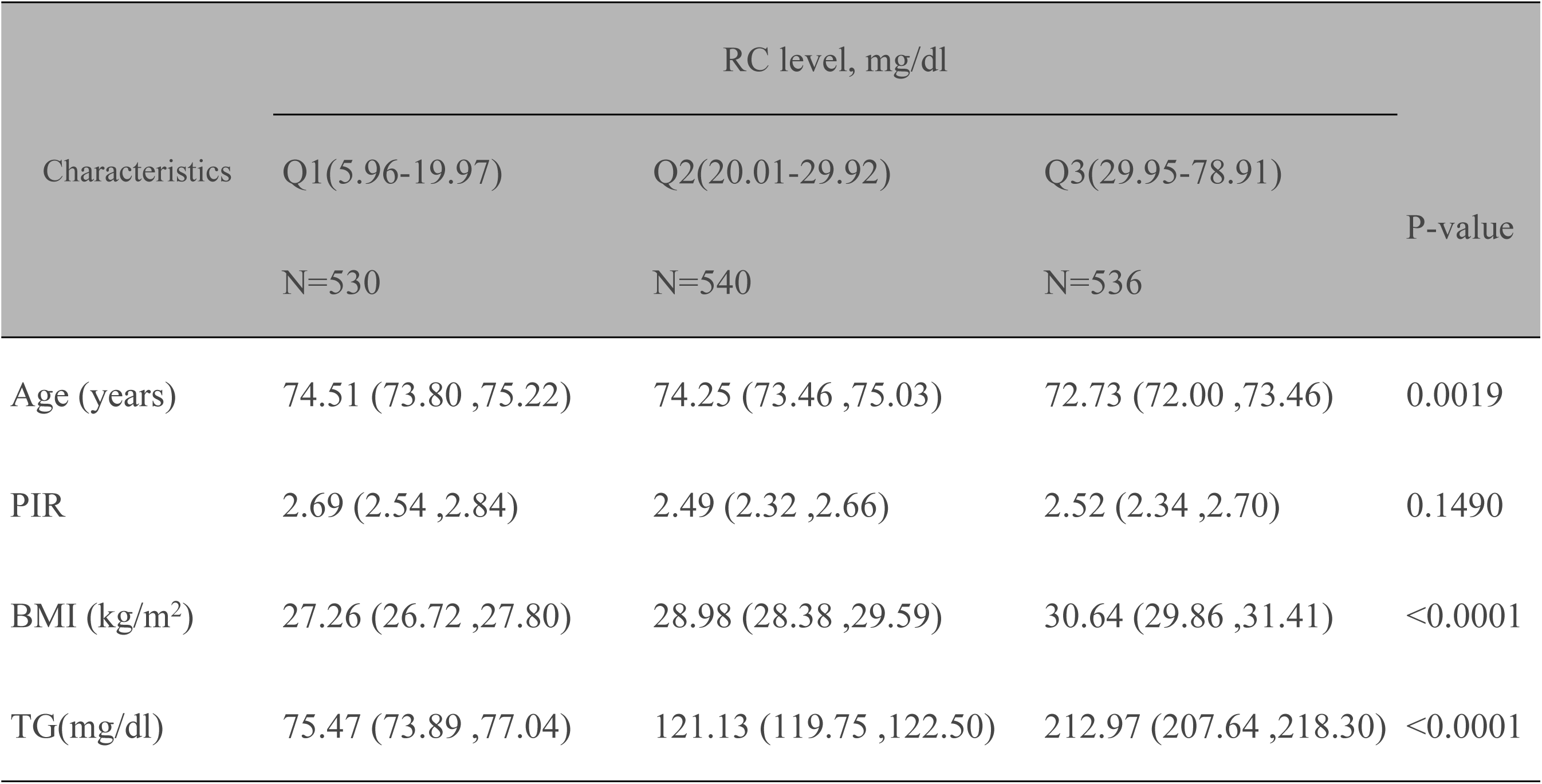
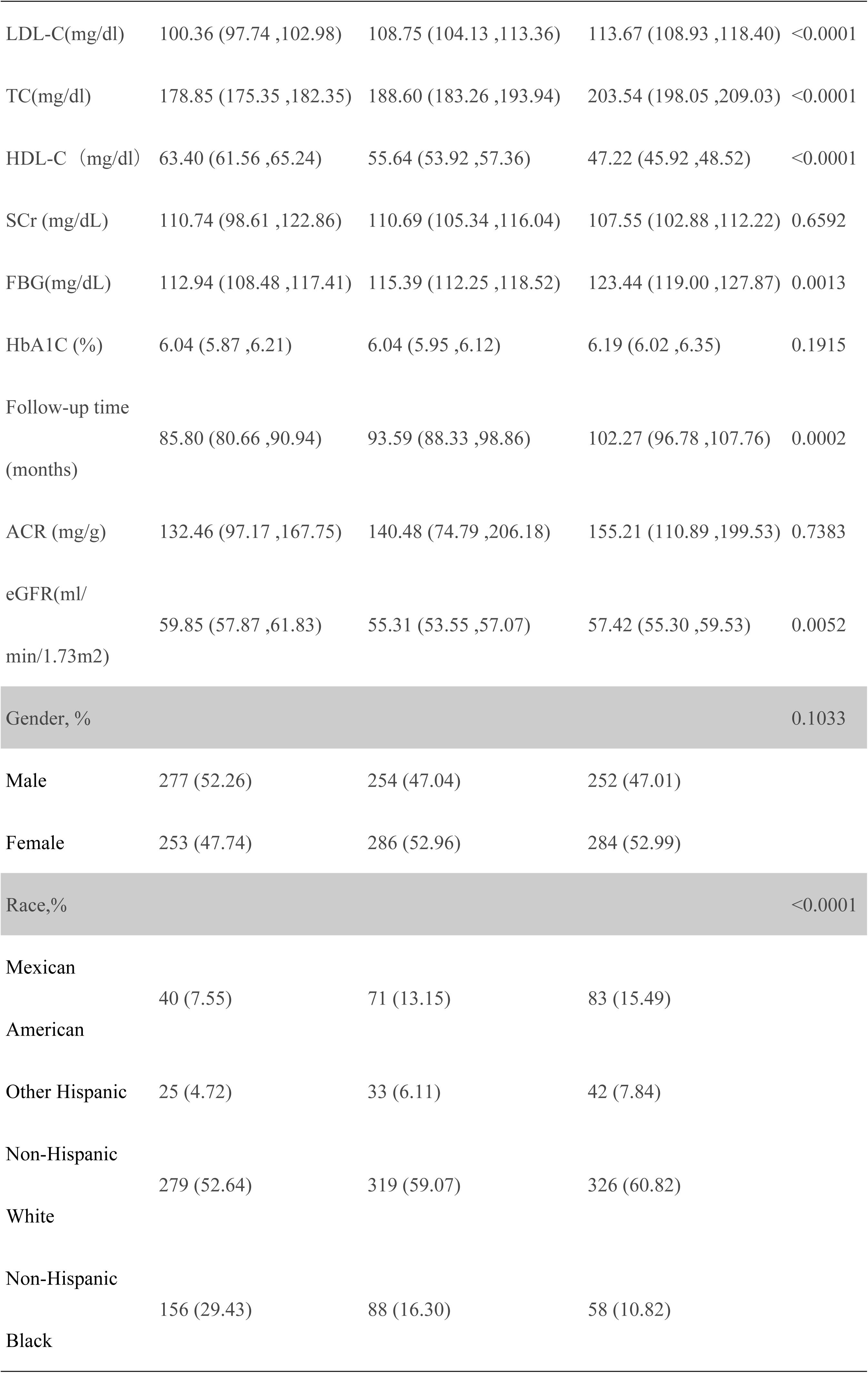
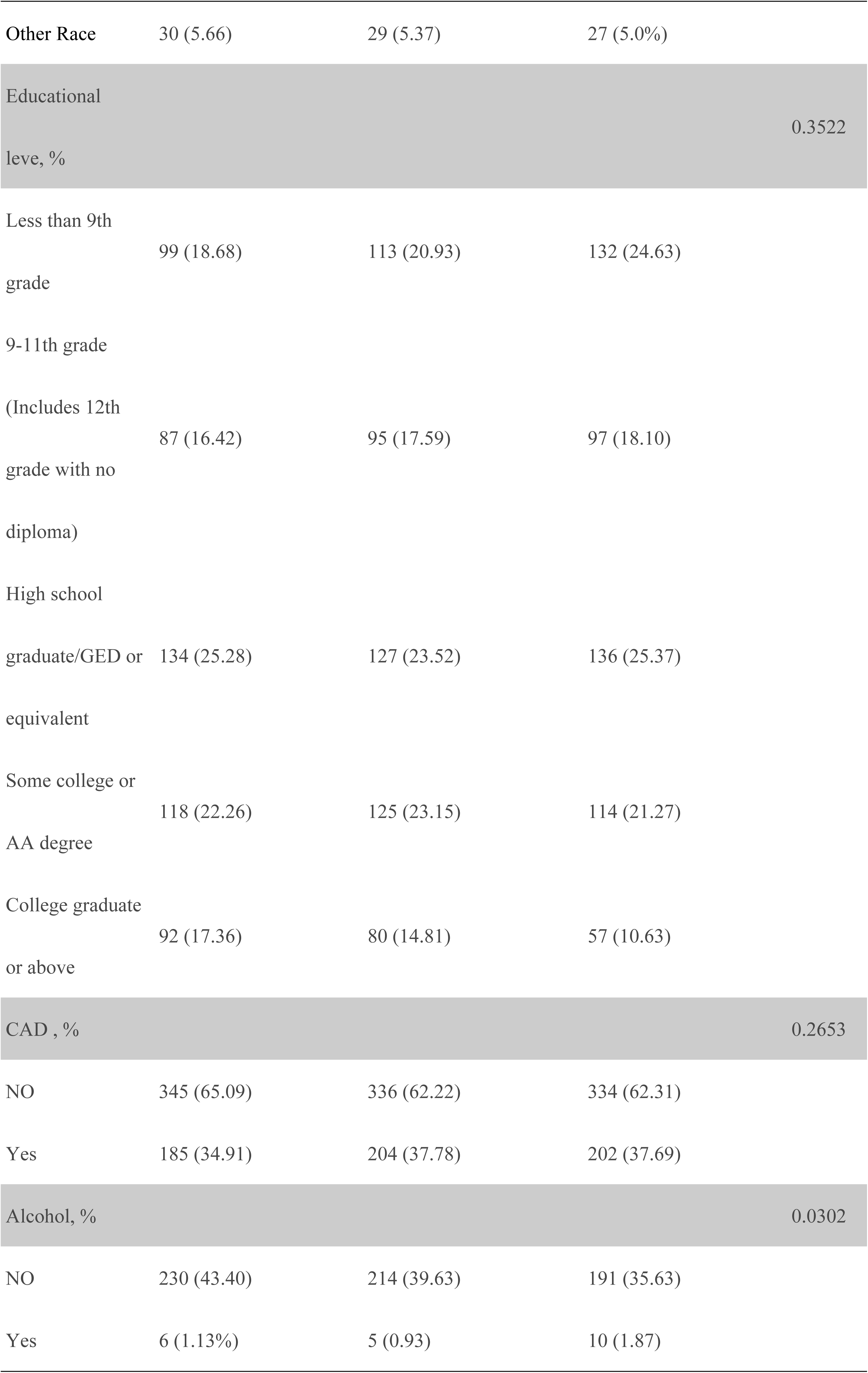
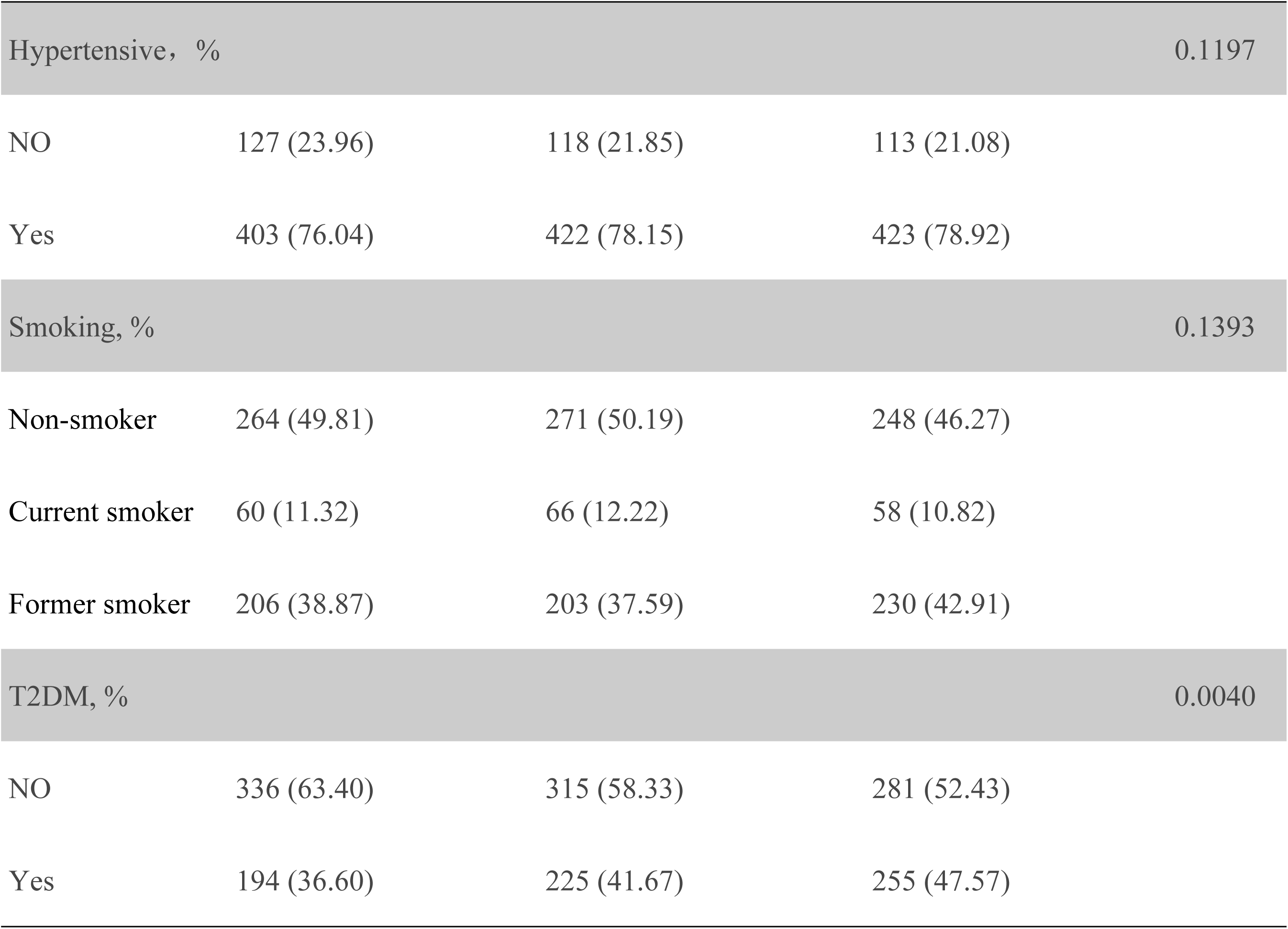
Baseline characteristics of study population according to RC(mg/dl) tertiles, weighted. Note: Continuous data: the mean with 95% confidence intervals, P value was calculated by weighted linear regression model. Categorical data: the number of occurrences with respective percentages, P value was calculated by weighted χ^2^ test. Abbreviations: PIR,Ratio of family income to poverty;BMI,body mass index;TG,Triglycerides;LDL-C, low-density lipoprotein cholesterol;Scr,Creatinine; TC,Total cholesterol;FBG,Fasting blood glucose;HbA1C,Hemoglobin A1c;CAD, coronary atherosclerotic heart disease; T2DM,diabetess;HDL-C, high-density lipoprotein-cholesterol;ACR,the urinary albumin to creatinine ratio;eGFR,estimated glomerular filtration rate.

## Results

### Baseline characteristics of study participants

In this stratified analysis of RC, significant demographic and clinical characteristic variations were observed across tertiles (5.96-19.97 mg/dL to 29.95-78.91 mg/dL), as shown in Table 1. The high RC group showed a lower mean age (72.73 vs. 74.51 years, P=0.0019) and increased body mass index (27.26 to 30.64 kg/m^2^, P<0.0001). Lipid profile analysis revealed escalating triglyceride levels (75.47 to 212.97 mg/dL, P<0.0001) and incrementally rising LDL-C (100.36 to 113.67 mg/dL, P<0.0001), while HDL-C significantly declined (63.40 to 47.22 mg/dL, P<0.0001). Glucose metabolism showed increased fasting blood glucose (112.94 to 123.44 mg/dL, P=0.0013) and higher Type 2 diabetes mellitus prevalence (36.60% to 47.57%, P=0.0040). Racial distribution demonstrated a significant association with RC levels (P<0.0001). Renal function assessment indicated slight but statistically significant eGFR differences (P=0.0052), with follow-up time increasing from 85.80 to 102.27 months (P=0.0002).

### Association of RC With All-Cause and CVD Mortality

In this study, we investigated the association between RC levels and mortality risks using Cox proportional hazards models with progressive covariate adjustments, as illustrated in Table 2. During the follow-up period of 93.9 months (approximately 7.8 years), a total of 932 all-cause mortality events were observed (representing 58.0% of the total population) and 283 cardiovascular mortality events (representing 17.6% of the total population).Our findings reveal a significant inverse relationship between RC and mortality, particularly noteworthy in all-cause mortality analysis. Specifically, for each logarithmic unit increase in RC, we observed a substantial 41% reduction in total mortality risk (adjusted HR: 0.59, 95% CI: 0.40-0.87, P=0.0079). Quartile-based analysis further substantiated this trend, demonstrating that the highest RC group (Q3) exhibited a 21% lower mortality risk compared to the lowest group (Q1, HR: 0.79, 95% CI: 0.65-0.96, P=0.0153), with a statistically significant dose-response relationship (P trend = 0.0163). Regarding cardiovascular mortality, while the results showed a promising 48% risk reduction (HR: 0.52, 95% CI: 0.27-1.015, P=0.0552), the findings did not reach conventional statistical significance. The quartile analysis of cardiovascular mortality mirrored this pattern, with the highest RC group showing a 26% risk reduction (HR: 0.74, 95% CI: 0.52-1.06, P=0.1018), though again without achieving statistical significance. Moreover, as shown in Fig. 2, generalized additive model analysis indicates that there is no nonlinear relationship between RC and total cause and cardiovascular mortality rates.

**Fig 2.**
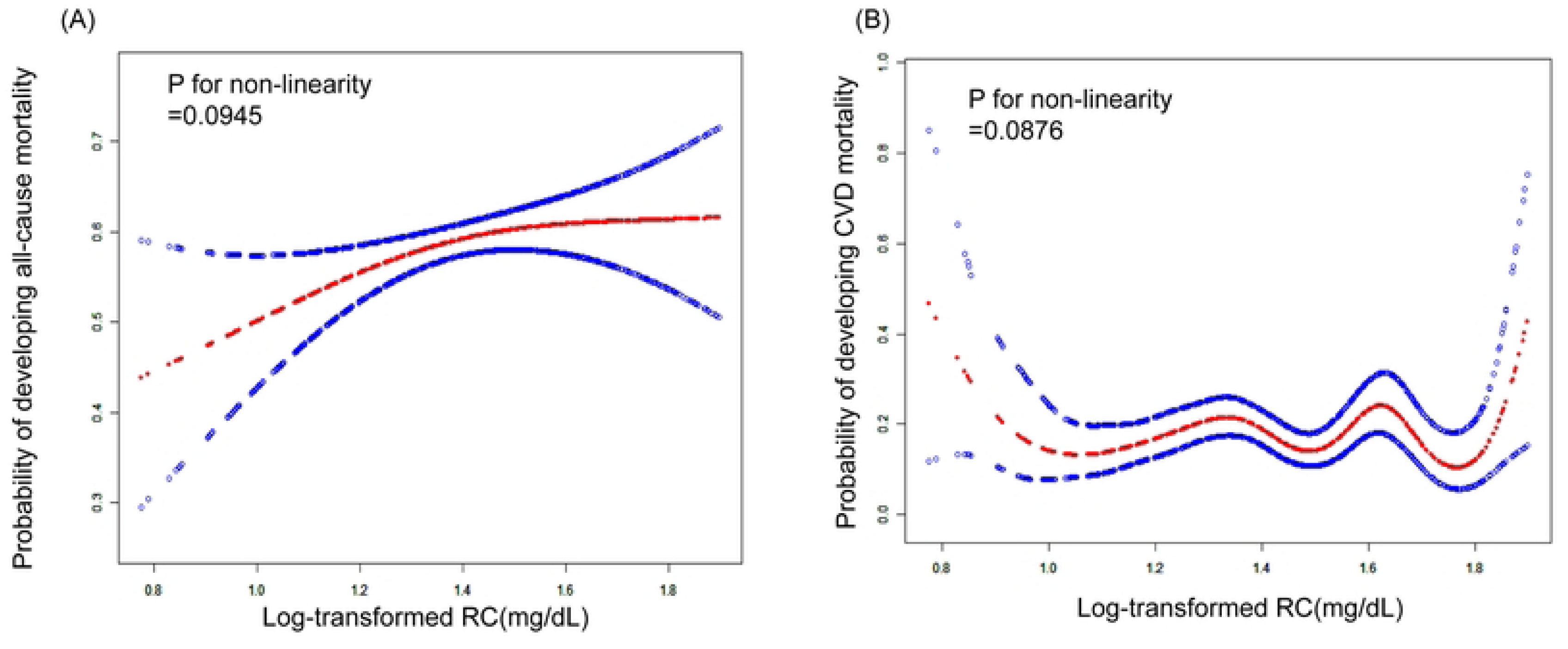
The generalized additive model analyses for the association between log-transformed RC and the risk of cardiovascular disease (CVD) and all-cause mortality. All covariates from model 3 were adjusted. (A) The relationship between log-transformed RC levels and all-cause mortality. The red line represents the estimated probability of all-cause mortality, and the blue dashed lines represent the 95% confidence intervals. The P-value for non-linearity was 0.0945, suggesting a potential non-linear association between RC levels and all-cause mortality risk, although not reaching statistical significance.(B) The relationship between log-transformed RC levels and cardiovascular disease (CVD) mortality. The red line indicates the estimated probability of CVD mortality, with blue dashed lines showing the 95% confidence intervals. The P-value for non-linearity was 0.0876, indicating a possible non-linear relationship between RC levels and CVD mortality risk, though not statistically significant.

**Table 2.**
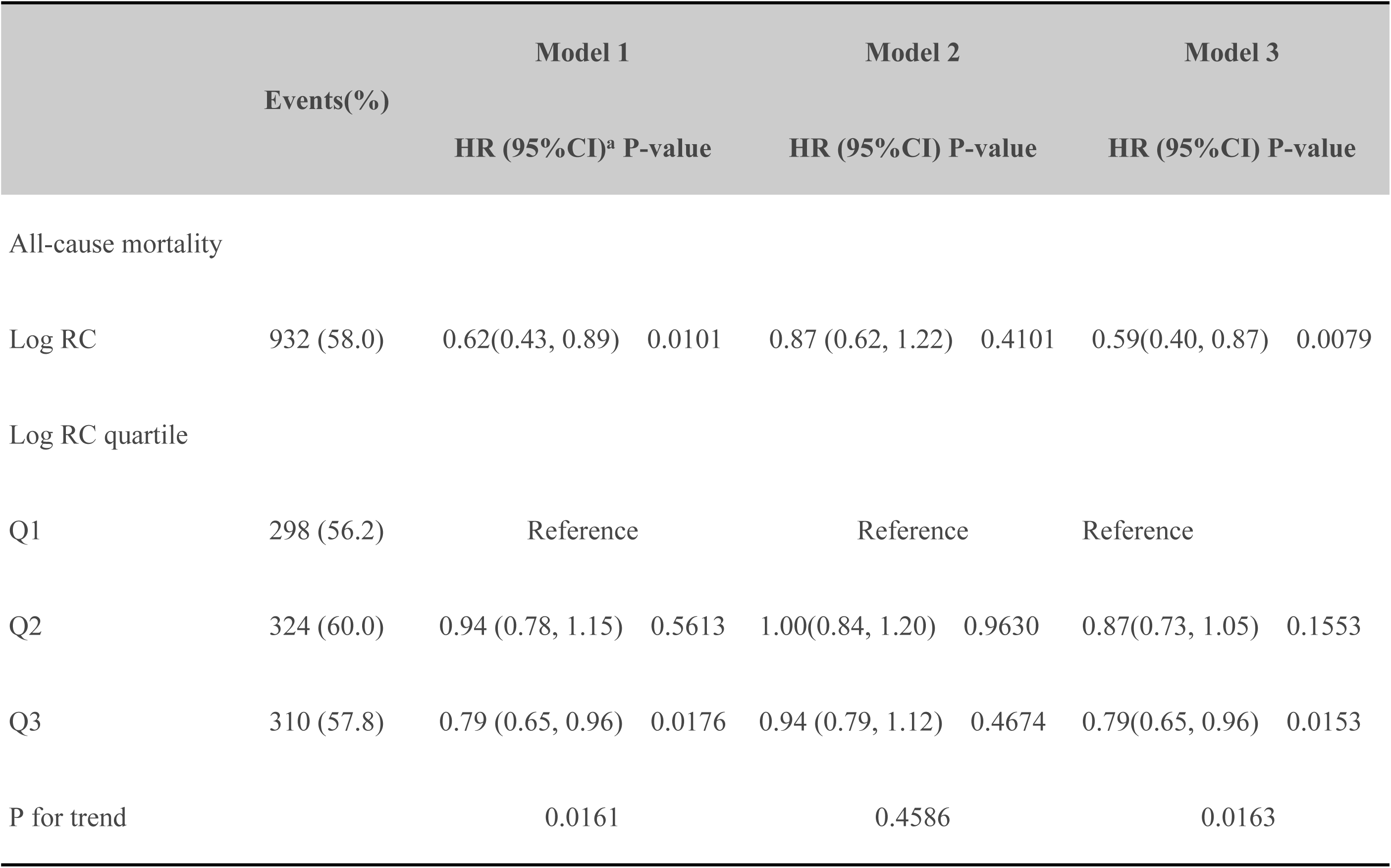
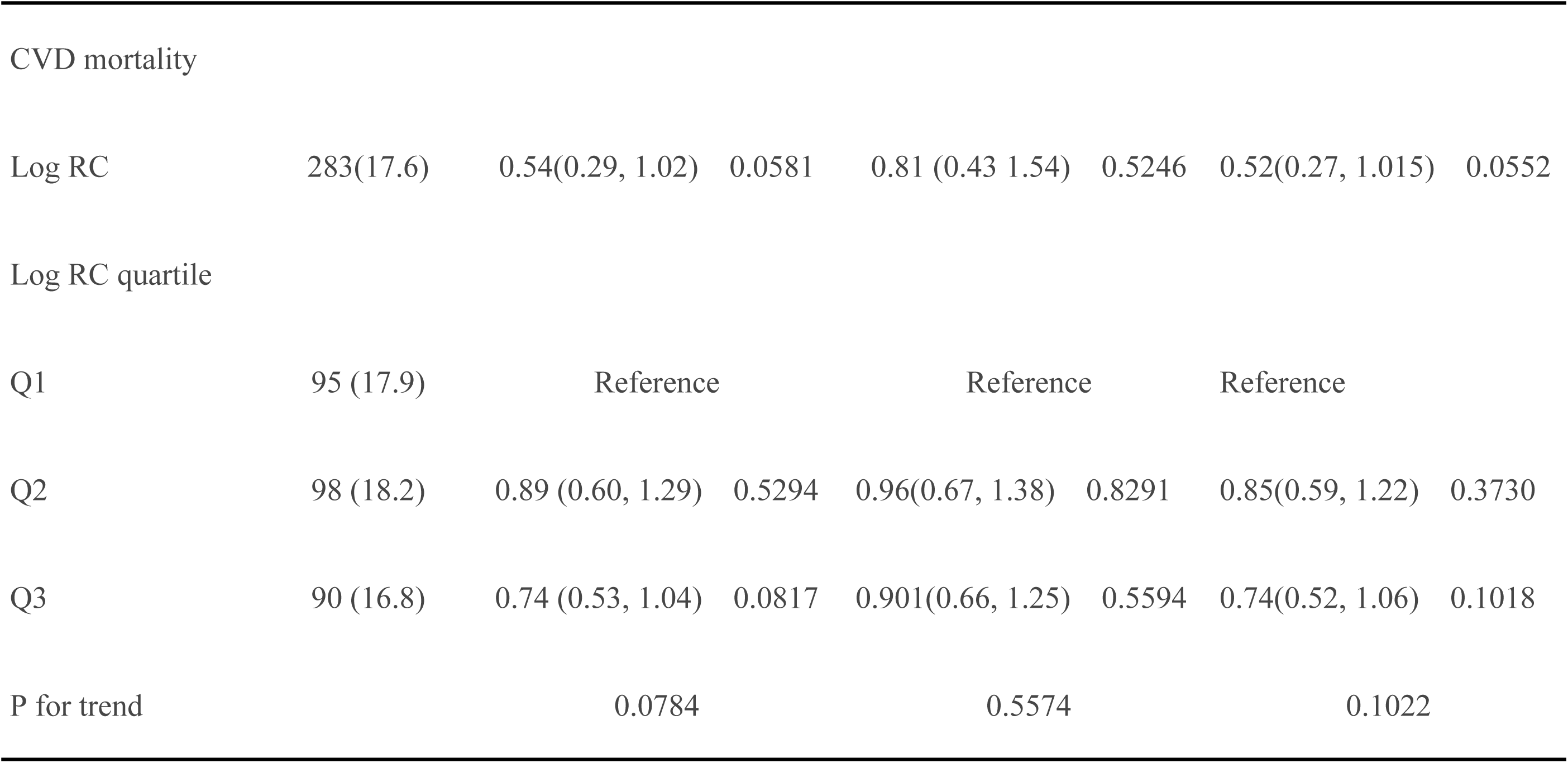
Association of log-transformed RC levels With All-Cause and CVD Mortality. Model 1: no covariates were adjusted. Model 2: age, gender, and race were adjusted. Model 3: age, gender, race,BMI,Hypertensive,SCr,smooking, Alcohol,FBG, CAD,T2DM,Education level and PIR were adjusted. PIR,Ratio of family income to poverty;BMI,body mass index;T2DM,diabetess;SCr,Creatinine; FBG,Fasting blood glucose;CAD, coronary atherosclerotic heart disease;survey-weighted percentage;HR, hazard ratio.Cox proportional hazards models were used to estimate hazard ratios (HRs) and 95% confidence intervals (CIs).

|  |  | Model 1 |  | Model 2 |  | Model 3 |  |
| --- | --- | --- | --- | --- | --- | --- | --- |
|  |  | HR (95%CI)<sup>a</sup> | P-value | HR (95%CI) | P-value | HR (95%CI) | P-value |
| All-cause mortality |  |  |  |  |  |  |  |
| Log RC | 932 (58.0) | 0.62(0.43, 0.89) | 0.0101 | 0.87 (0.62, 1.22) | 0.4101 | 0.59(0.40, 0.87) | 0.0079 |
| Log RC quartile |  |  |  |  |  |  |  |
| Q1 | 298 (56.2) | Reference |  | Reference |  | Reference |  |
| Q2 | 324 (60.0) | 0.94 (0.78, 1.15) | 0.5613 | 1.00(0.84, 1.20) | 0.9630 | 0.87(0.73, 1.05) | 0.1553 |
| Q3 | 310 (57.8) | 0.79 (0.65, 0.96) | 0.0176 | 0.94 (0.79, 1.12) | 0.4674 | 0.79(0.65, 0.96) | 0.0153 |
| P for trend |  | 0.0161 |  | 0.4586 |  | 0.0163 |  |
| CVD mortality |  |  |  |  |  |  |  |
| Log RC | 283(17.6) | 0.54(0.29, 1.02) | 0.0581 | 0.81 (0.43 1.54) | 0.5246 | 0.52(0.27, 1.015) | 0.0552 |
| Log RC quartile |  |  |  |  |  |  |  |
| Q1 | 95 (17.9) | Reference |  | Reference |  | Reference |  |
| Q2 | 98 (18.2) | 0.89 (0.60, 1.29) | 0.5294 | 0.96(0.67, 1.38) | 0.8291 | 0.85(0.59, 1.22) | 0.3730 |
| Q3 | 90 (16.8) | 0.74 (0.53, 1.04) | 0.0817 | 0.901(0.66, 1.25) | 0.5594 | 0.74(0.52, 1.06) | 0.1018 |
| P for trend |  | 0.0784 |  | 0.5574 |  | 0.1022 |  |

### Subgroup analysis

As shown in Table 3, after adjusting for confounding factors in Model 3, we observed significant associations in this comprehensive stratified analysis of residual cholesterol (RC) and mortality risk within specific subgroups. Women (HR=0.71, 95% CI: 0.54-0.93, P=0.0132), patients with hypertension (HR=0.75, 95% CI: 0.59-0.95, P=0.0151), and patients with type 2 diabetes (HR=0.66, 95% CI: 0.48-0.91, P=0.0111) exhibited statistically significant reductions in all-cause mortality risk. Notably, non-smokers also demonstrated a significant decrease in cardiovascular mortality (HR=0.57, 95% CI: 0.33-0.99, P=0.0449). Although differences among subgroups were observed, the interaction P-values (all >0.05) indicate that the effects of RC are consistent across different population strata, demonstrating that the strong association between RC and mortality risk remains relatively stable across various demographic and clinical characteristics.

**Table 3.**
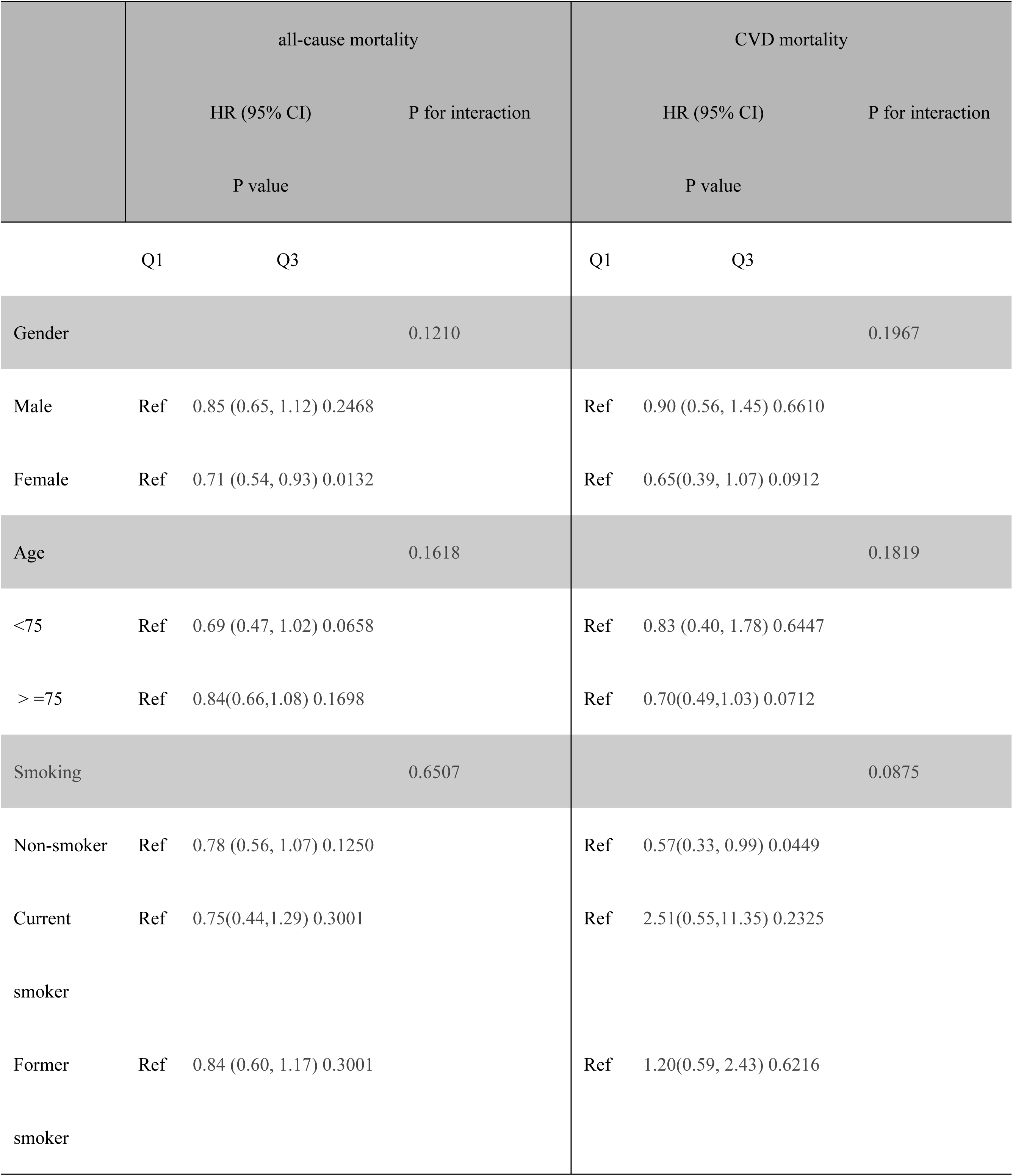
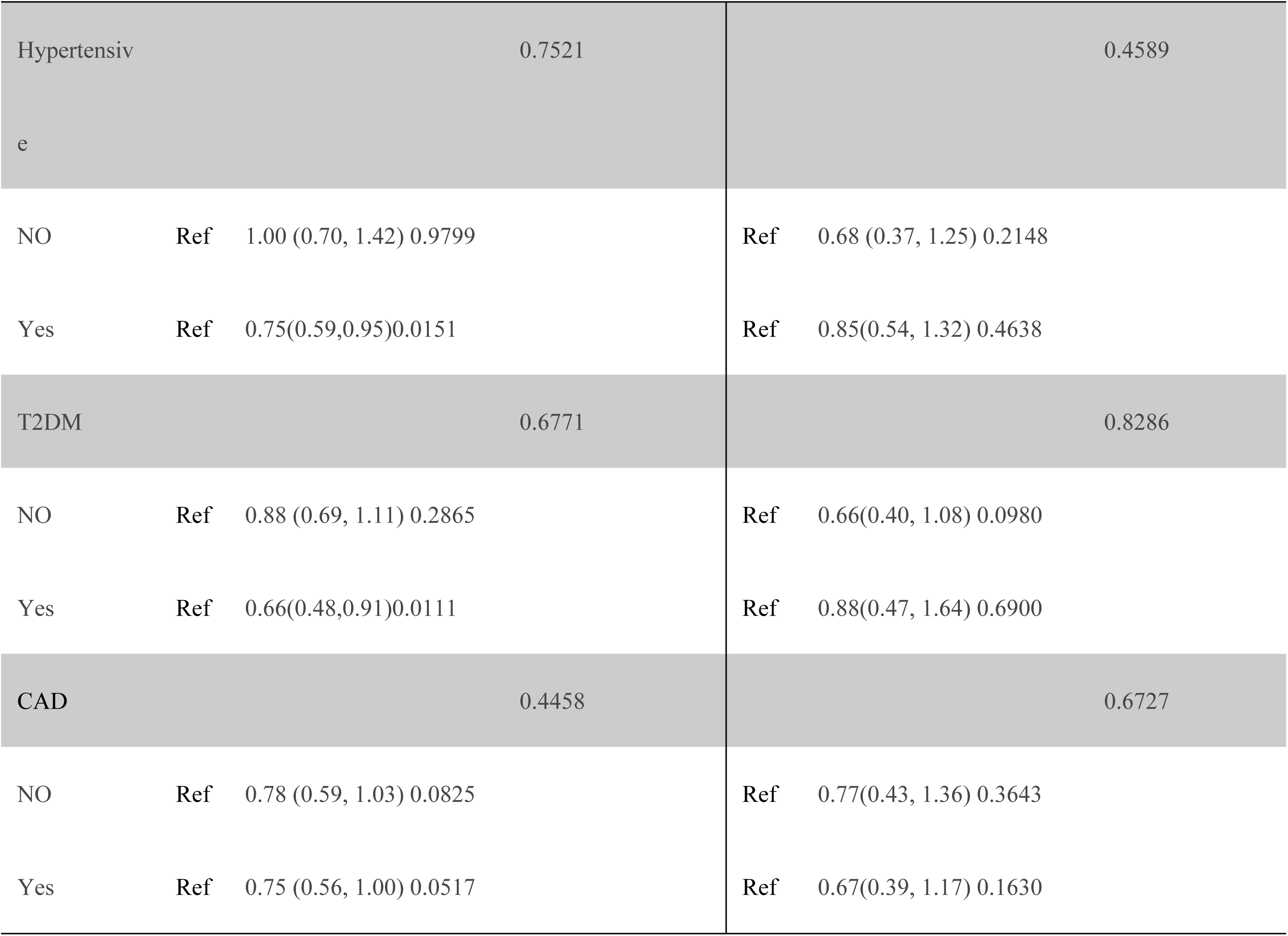
Stratified analyses of the associations between log-transformed RC levels and all-cause mortality and CVD mortality. Note:CAD, coronary atherosclerotic heart disease; T2DM,diabetess;HR, Hazard ratio; CI, confidence interval; Ref, reference. survey-weighted percentage.age, gender, race,BMI,Hypertensive,SCr,smooking, Alcohol,FBG, CAD,T2DM,Education level and PIR were adjusted. PIR,Ratio of family income to poverty;BMI,body mass index;T2DM,diabetess;SCr,Creatinine; FBG,Fasting blood glucose;CAD, coronary atherosclerotic heart disease;Cox proportional hazards models were used to estimate HRs and 95% 95% CIs.

## Discussion

In this NHANES-based cohort study (2001–2018), we examined the association between RC and mortality in CKD patients over 60. Among 1,606 participants followed for 7.8 years, 932 all-cause and 283 cardiovascular mortality events occurred. Notably, each logarithmic RC unit increase was linked to a 41% reduction in all-cause mortality risk (adjusted HR: 0.59, 95% CI: 0.40-0.87, P=0.0079), with the highest RC group showing a 21% lower mortality risk compared to the lowest group (HR: 0.79, 95% CI: 0.65-0.96, P=0.0153).

Our study revealed a significant negative correlation between RC and all-cause mortality risk in CKD patients, partially consistent with the findings of Yu et al.(11) published in the Journal of Clinical Endocrinology & Metabolism in 2021. Yu’s study also focused on patients with type 2 diabetes and nephropathy, discovering a complex association between RC and mortality risk. However, our study offers notable methodological advantages: first, we utilized a larger NHANES database (1,606 patients) with a follow-up period of 93.9 months, compared to the relatively smaller sample in Yu’s research. Second, our study population comprised CKD patients over 60, providing a more representative sample and reducing potential bias through multi-model adjustments. Notably, we observed a 41% reduction in all-cause mortality risk for each logarithmic unit increase in RC, a finding that subtly differs from Zhao et al.’s(12) research in Frontiers in Cardiovascular Medicine on heart failure patients. While Zhao’s study highlighted RC’s predictive value for mortality in heart failure patients, our research unveiled a protective effect of RC in CKD patients. These differences may stem from metabolic characteristics and inflammatory states across different disease populations. For instance, Yuan et al.’s(8)research indicates that RC is closely associated with the pre-inflammatory state in kidney disease patients, potentially explaining the observed association between RC and reduced mortality risk. Our subgroup analysis further revealed that the mortality risk-reducing effect of RC was more pronounced in women, hypertensive, and type 2 diabetes patients, providing crucial insights for personalized medicine.Furthermore,the intricate physiological mechanisms of RC in CKD patients unveil its complex association with mortality risk. Hua et al. elucidated the intricate interplay between inflammation and lipid metabolism in CKD, revealing the multifaceted metabolic regulatory pathways of RC(27). Chen et al. investigated the mechanistic role of remnant lipoproteins in cardiovascular disease, demonstrating that RC transcends its traditional role as a mere lipid transport molecule, emerging as a critical modulator of inflammatory and metabolic processes(28). Butcko et al. emphasized that inflammatory responses in CKD patients represent a pivotal perspective for comprehending the physiological implications of RC(29). Our study reveals a non-linear relationship between RC and mortality risk, arising from its nuanced regulatory functions in inflammation, energy metabolism, and vascular homeostasis, thereby providing a novel theoretical framework for personalized risk stratification.

Through an in-depth exploration of RC and mortality risk in CKD patients, our study offers a crucial new clinical risk assessment perspective. We first systematically revealed the inverse association between RC and all-cause mortality in a large population, challenging traditional risk perceptions. The significant RC-related mortality risk reduction in women, hypertensive, and type 2 diabetes subgroups provides critical guidance for targeted interventions. Future research should investigate RC’s biological mechanisms and differential impacts across chronic disease populations, developing risk stratification and personalized prediction models. We recommend incorporating RC into routine lipid profile and mortality risk assessments, using multi-dimensional tracking to enhance individualized medical management for CKD patients.

The study’s strengths lie in its rigorous methodological design and comprehensive data analysis. Utilizing the NHANES database with complex stratified multi-stage sampling, we ensured high representativeness and scientific rigor. The research included 1,606 CKD patients over 60, with a 7.8-year follow-up, providing a stable observation window for exploring RC and mortality risk. We employed a multi-layered Cox proportional hazards model, progressively adjusting for potential confounding factors like age, gender, race, BMI, hypertension, smoking, and diabetes to minimize bias. Innovatively using generalized additive models and penalized spline methods, we explored the dose-response relationship between RC and mortality risk. Subgroup analysis revealed RC’s differential impacts across population characteristics, uniquely demonstrating its significant role in reducing mortality risk among women, hypertensive, and type 2 diabetes patients, offering new theoretical and practical insights for personalized medicine.

While providing important insights into RC and mortality risk in CKD patients, our study has notable limitations. Based on NHANES data focusing on American patients over 60, our findings may have limited generalizability across different races, age groups, and geographical regions. As an observational cohort study, we can only demonstrate an association, not establish a causal mechanism. Despite adjusting for known confounders, potential unmeasured factors like genetic background and lifestyle details may influence result accuracy. The 7.8-year follow-up may not fully capture the long-term impact of RC dynamics on mortality risk. We did not extensively explore the specific biological mechanisms underlying this association, providing a crucial direction for future research. We recommend subsequent multi-center, large-sample prospective studies combined with molecular biological investigations to further validate and expand our findings.

## Conclusions

This study reveals a novel association between RC and mortality risk in CKD patients. Our findings suggest RC as a potential prognostic indicator, expanding understanding of its physiological role and offering new perspectives for personalized clinical management.

## Data Availability

All data are freely available through the NHANES database (https://www.cdc.gov/nchs/nhanes/)

## Abbreviations

PIR: Ratio of family income to poverty
BMI: Body mass index
TG: Triglycerides
LDL-C: Low-density lipoprotein
TC: Total cholesterol
HDL-C: High-density lipoprotein-cholesterol
SCr: Creatinine
FBG: Fasting blood glucose
HbA1C: Hemoglobin A1c
CAD: Coronary atherosclerotic heart disease
T2DM: Diabetess
CKD: Chronic kidney disease
RC: Remnant cholesterol
NHANES: Nutrition Examination Survey

## Data Availability

All data are freely available through the NHANES database (https://www.cdc.gov/nchs/nhanes/)

## Author contributions

J.C.Y. conceived and designed the study, and wrote the manuscript. C.L. analyzed the data. Q.B.X. took the quality control of data. G.D.L. and Y.L. critically revised the manuscript. All authors read and approved the final manuscript.

